# Biological diagnosis and characterization of occult hepatitis B virus infection in Burkina Faso

**DOI:** 10.1101/2023.01.04.23284178

**Authors:** Boubacar Doumbia, Birama Diarra, Bourahima Kone, Florencia Djigma, Bocar Baya, Jacque Simpore

## Abstract

**Introduction/objective:** The occult infection by the hepatitis B virus (OBI) characterized by the undetectable (HBs Ag) negative antigen in the serum and the presence of viral DNA, constitutes a major public health problem and a major challenge for clinical forms of hepatitis worldwide. The persistence of OBI very often leads to hepatocellular carcinoma (HCC) and immunodeficiency. The aim of this study was to estimate the prevalence of OBI and to characterize the incriminated genotypes of the virus.

**Material and method:** The plasmas of 252 HBsAg-negative volunteers were used for highlighting the viral DNA by PCR classic. PCR Multiplex using specific primers of 6 genotypes (A to F) was used for characterization.

**Results:** A prevalence of 11.9% (30/252) of OBI was obtained with 7.5% in women and 4.4% for men. Genotypes E (60.0%) and A3 (23.3%) were present and genotypes B, C, D, and F were absent. A mixed infection with E/A3 genotypes (16.7%) was observed. AC Anti-HBc was present in 80% of cases OBI.

**Discussion/Conclusion:** The prevalence of 11.9% of OBI could be related to the level of endemicity of the study area, Burkina Faso being a country with a prevalence of more than 14% of HBV infection. These infections are dominated by the A3 and E genotypes, confirming their importance in chronic HBV infections. The absence of genotypes B and C in cases of OBI constitutes a positive predictive value since their presence is linked to a more rapid evolution of the infection towards HCC.

## 1. Introduction

Hepatitis B virus (HBV) infection remains a major public health problem in the world. According to the World Health Organization, approximately 2 billion people are infected with the hepatitis B virus (HBV). Carriers of chronic hepatitis B are estimated at about 350 million, of whom more than 700,000 die each year from cirrhosis and/or hepatocellular carcinoma (HCC) [1]. Burkina Faso (BF) is a highly endemic country with HBV prevalence varying between 8% and 14.5% in the general population [2, 3]. Prevalences of 14.3%; 17% and 12.9% were reported respectively among blood donors from Nouna, from Ouagadougou and the national blood transfusion center of Burkina Faso [4, 5]. Blood transfusion constitutes of a major risk factor for transmission of the OBI virus following blood screening of donors without the use of nucleic amplification tests [6, 7]. The use of nucleic acid amplification tests to screen blood donors for HCV, HIV, HBV, and OBI can prevent the occurrence of infections due to these viruses [8]. The implementation of these nucleic amplification tests is not yet an achievement for developing countries. The prevalence of OBI among blood donors varies from country to country [9]. In Burkina Faso, this prevalence was around 32.8% among blood donors [10]. The detection of HBV surface antigen (HBsAg) in serum remains the basic element in the diagnosis of chronic infection and for HBV screening in developing countries [11]. And the majority of HBsAg positive individuals are also positive for HBV DNA in serum. Occult HBV infection (OBI) is characterized by the presence of viral DNA in individuals with undetectable HBsAg, with the presence or absence of anti-HBc antibody [12]. Chronic carriers of hepatitis C virus (HCV) and human immunodeficiency virus (HIV) are associated with a high prevalence of OBI [13, 14]. OBI/HCV coinfection accelerates the progression of hepatitis to cirrhosis and increases the risk of progression to hepatocellular carcinoma (HCC) [15]. This coinfection also decreases the response to hepatitis treatment with the alpha interferon [16]. The molecular mechanisms underlying the occurrence of OBI play a direct role in the development of hepatocellular carcinoma (HCC) [17, 18]. The HBV genome is genetically variable. Phylogenetic studies have classified HBV into 10 genotypes (A-J) [19, 20]. These genotypes and their variants (sub-genotypes) have a distinct geographic distribution and have been associated with the development of cirrhosis and liver cancer (HCC) [21]. This genetic variability of HBV plays an important role in the occurrence of OBI [22, 23]. In Burkina Faso, we have few data on the prevalence of occult hepatitis B virus infection in the general population. The aim of this study was to estimate the prevalence of occult HBV infection and to characterize the genotypes of the virus implicated in volunteers in the population of Ouagadougou.

## 2. Materials and Methods

### Study framework

The study took place at the laboratory of Biological and Molecular Genetic (LAOBIGENE) of the University Ouaga I Pr Joseph Ki-Zerbo in Burkina Faso.

### Type, populations and study period

This is a prospective study, which was carried out between June 2018 and December 2019. This is a population of two hundred and fifty-two (252) volunteers residing in Ouagadougou, without distinction of age, professions or social categories who have given their free and informed consent. For children, consents have been obtained from parents or guardians. The recruitment of volunteers took place in different sentinel sites in Ouagadougou after an awareness campaign lasting several days for screening for hepatitis B. After obtaining the free and informed consent of the participants, screening for HBV was carried out in using a rapid diagnostic test (RDT). The collection of blood samples concerned the HBs antigen negative participants.

### Sampling

A blood sample was taken from all participants who were HBs antigen negative. A volume of 4 ml of venous blood was drawn into the EDTA tubes. After sedimentation for two hours (2 hours), the plasmas were decanted from the pellets to prepare different aliquots. These plasma aliquots were stored at −20°C in freezers before proceeding to HBV detection.

### Hepatitis B virus (HBV) detection

#### ➢DNA extraction

It was performed using the Genomic Column DNA Express kit (Sacace Biotechnologies, Como, Italy) according to the manufacturer’s protocol. The protocol uses a column extraction method based on a principle of retention of nucleic acid molecules on a silicate membrane contained in a column. The cells are first lysed by a solution containing chaotropic ions in the presence of proteinase K. After a washing step which rids the sample of contaminants, the nucleic acid is recovered by elution in a slightly alkaline solution. The pure product (the nucleic acid) can therefore be directly amplified.

#### ➢Classical PCR

It was carried out using the procedure of the manufacturer AmpliTaq Gold 360 Master Mix, by amplification of a very conserved region (Pres-S) of 1104 bp of the HBV genome. The primer pair used (Table I) was designed from a highly conserved region of the HBV S surface envelope. The PCR was carried out with the GeneAmp PCR System 9700 ® device (Applied Biosystems, USA) in a reaction volume of 25 μl which contained 12.5 μl of Master mix Ampli Gold Taq Man (Applied Biosystems, USA), 1 μl of forward and reverse primers and 5.5 µl of sterile water and 5 µl of DNA. The PCR products were subjected to electrophoresis on a 2% agarose gel prepared with ethidium bromide (10 mg/ml), and visualized under UV light at 312 nm.

#### Electrophoresis and Revelation under Ultra Violet (UV) light

The transparent support containing the gel was placed in the electrophoresis tank containing migration buffer. The samples were placed on the gel while ensuring that the migration buffer previously poured into the tank covers the gel by a few millimeters. To do this, 5 μl of loading blue were mixed with 5 μl of amplified cDNA per sample and placed in appropriate wells on the gel. Three (3) μl of the molecular weight marker (Ladder) were deposited in the first well. Electrophoretic migration at 120 V for 45 minutes. The gel was revealed under UV at 312 nm then photographed by the “GENE FLASH®” device. Results interpretation: The PCR was declared positive if the sample shows a band corresponding to the size of the expected DNA fragment **(Table I)**. It was negative in the absence of band.

#### ➢Characterization of genotypes by multiplex PCR

The multiplex PCR was carried out using the method described by Chen et al, with the following modifications: Two PCR-Multiplexes under the same conditions with two Mixes. Mix 1 will characterize genotypes A to C and Mix 2 genotypes D to F. The two PCR-Multiplexes were carried out in a total volume of 25 μl containing 12.5 μl of Master mix Ampli Gold Taq Man® (Applied Biosystems, USA), 1 μl of sense and antisense primers of the 3 genotypes **(Table II)**, 1.5 µl of sterile water and 5 µl of DNA. Interpretation of results, the PCR was considered positive if the sample shows a band corresponding to the size of the expected DNA fragment. It was negative in the absence of the band. The sizes of the fragments and their corresponding genotypes are shown in Table II.

#### ➢Rapid detection tests for HBV markers Principles

##### HBs Ag and HBe Ag antigens

Immuno-chromatographic method using the ELISA technics (Enzyme Link Immuno-Sorbent Assay) in sandwich. The anti-HBsAg and anti-HBeAg antibodies fixed on the test membrane react with the HBsAg and HBeAg antigens of the serum to be tested and the Ag-AC couples migrate by capillary action to encounter other anti-HBsAg and anti-Ag antibodies at the second level of the test. HBe and give a colored reaction indicating the positivity of the test.

##### Anti-HBs antibodies

Immuno-chromatographic method, it consists of fixing the anti-HBs antibodies of serum by HBsAg previously fixed to the membrane of the test, then fixing by a second sandwiched HBsAg at the second level of the test after migration by capillarity.

##### Anti HBe and anti HBC antibodies

Immuno-chromatographic method using the competitive ELISA technique. The anti-HBe and anti-HBc antibodies compete with other anti-HBe and anti-HBc antibodies fixed on the membrane, for the HBeAg and HBcAg previously fixed on the membrane. The positive result is characterized by the absence of a line because there is competition between the antibodies.

##### Data collection tools

The data was entered on Excel software and analyzed on SPSS version 25 software. The significance level was set at p<0.05.

##### Ethical consideration

This study was carried out taking into account the free and informed consent of the participants. Respect for confidentiality and anonymity in relation to the provided information was essential. The study caused the approval of the institutional ethic committee of CERBA/LAOBIGENE and the national ethics committee of the Ministry of Health of Burkina Faso.

## 3. Results

The present study focused on 252 volunteers screened for viral hepatitis B virus (HBV), all negative for HBV surface antigen (HBsAg). Of the 252 individuals with undetectable HBsAg, viral DNA was present at 30 or 11.9%. The seroprevalence of occult HBV infection was 13.77% in women against 9.65% in men. It was respectively 9.30%, 16.39%, 3.39% and 14.29%, in the age groups of less than 20 years, 20-34 years, 35-50 years and more than 50 years. It was 10% based on rural origin against 11.98% urban origin. On the marital level, the prevalence was 12.50% and 11.43% respectively among singles and married people (**Table III**). The presence of genotypes E and A3 was recorded with respectively 60.0% (18/30) and 23.3% (7/30) (**Figure 1) (Table IV)**. Genotypes B, C, D, and F were absent. A mixed infection with E/A3 genotypes (5/30 or 16.7%) was identified in some samples. The prevalences of serological markers of occult HBV infection were respectively 80%, 13.3%, 10.0% and 0.0% for Ac HBc, Ac HBe, Ac HBs and Ag HBe respectively. The HBc Ac seropositive IOB was 80.0% against 20% of the seronegative IOB. No coinfection with HCV was found in the cases of IOB **(Table V)**.

**Image 1:**
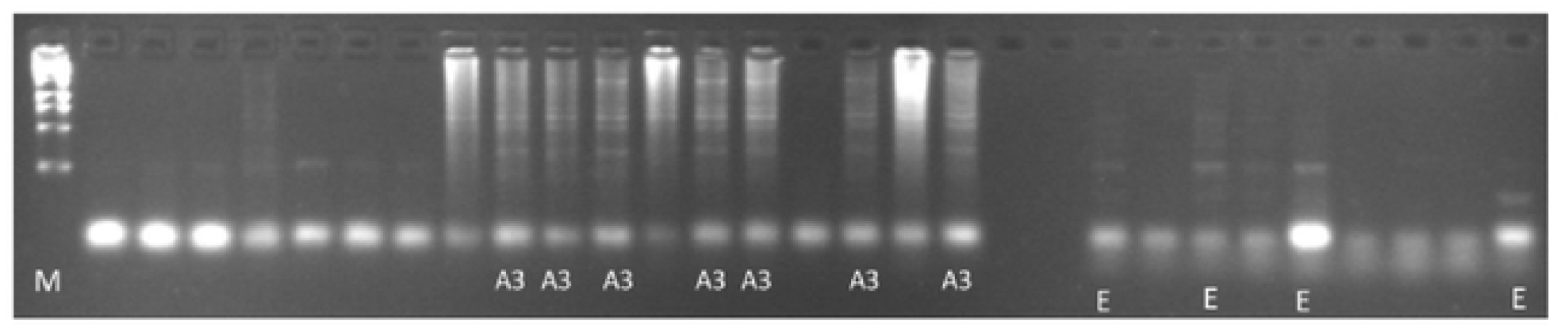
Photo of 2% agarose gel of genotypes E and A3.

## 4. Discussion

Occult hepatitis B defined by the presence of viral DNA in the serum and/or in the hepatocytes and the absence of detectable HBsAg, has been described since the 1970s. But during the last decades that it has grown the size because of the reinforcement of the measures of transfusional security and the organ transplantation development mostly in developed countries. Data on OBI in developing countries are almost non-existent. In the literature the prevalence of OBI varies according to the methods of detection and areas endemicity level. [9, 24]. Burkina Faso is a country with a high endemicity for hepatitis viral B with a prevalence of more than 14% [2].

Among the 252 individuals who make up our study population, 138 (54.8%) were female and 114 (45.2%) males. The sex ratio was 0.97 in the favor of women as was the case in the study conducted by Oluyinka and al, in Nigeria on OBI in blood donors, where the sex ratio was 0.62 [24]. However, in 2014 in Burkina Faso, Somda, found a sex ratio of 4 in the favor of men in their study [10]. The majority participation of women in our study can be explained by a much greater availability on their part in voluntary screening. The 20-34 age group was the largest in terms of the number of participants in our study, as was the case in the study conducted by Oluyinka and al. However, the 30-60 age group was the most important in the studies conducted by Kalantari and al, and Baghbanian and al, on the prevalence of OBI in hemodialysis patients and cancer patients in Iran [25, 26].

The distribution of OBI generally varies from region to region and according to risk group [9]. In the present study, we recorded a global prevalence of 11.9% (30/252) of OBI which may be related to the level of endemicity of the study of area to HBV infection. The prevalence of occult HBV infection varies according to the study of area and the study population, thus a prevalence of 17% (i.e. 72/429) was registered by Oluyinka and al., among Nigerian blood donors; 32.8% (25/76) was obtained by Somda and al., among Burkinabe blood donors; 5.5% (55/1000) by Shuaibu and al., among blood donors Malaysian; 0.13% (5/2972) by Hong Lin and al., among Chinese blood donors; 4.8% and 4.3% by Baghbanian and al., among Iran in blood and liver cancer patients respectively [9]. According to Makvandi and al., this variability in OBI prevalence depends on the sensitivity of the molecular techniques used (nested PCR, real-time PCR, or classical PCR) for the detection of viral DNA in serum and/or in hepatocytes, the size of the sample used and the geographic distribution of the study site. Thus, the prevalence of OBI varies from 1% (Canada) to 87% (Mexico) in different regions of the world [27]. However, these prevalences should be interpreted with great caution. Indeed, according to Samal and al., several factors can potentially influence the OBI estimation rate: risk groups (chronic HCV infection, HIV infection, cirrhosis, injection drug users, hemodialysis patients, and hepatocellular carcinoma), sample size, assays used for detection of mutant HBsAg and HBV genome amplification targets, high and low endemicity regions [11]. However, even in areas of low HBV endemicity such as Canada, cases of OBI have been reported [27]. The seroprevalence of OBI varied in the different groups that constituted the study population. Thus, this prevalence was 13.77% among women against 9.65% among men. According to age it was 9.30% among those under 20 and 3.39% between 35-50 years old. However, it had reached 16.39% among subjects aged 20 to 34 and 14.29 % in the over 50s. This variation of OBI in specific groups has also been recorded by other authors. Thus, Baghbanian and al., registered a prevalence of 4.3% in solid cancer patients and 4.8% in blood cancer patients [25]. There was little difference between the prevalence in rural areas 10% and that in urban areas 11.98% this similarity can be explained by the fact that it was the same study area. Characterization of genotypes currently the genotypes found in cases of OBI in different regions of the world are exclusively A, C, G, E and D [9]. In the present study we registered two genotypes E and A3. These A3 and E genotypes were prevalent with respectively 23.3% and 60%, similar results were found by Kurbanoy and al., in 2005 in Cameroon where genotypes A and E were in the majority with 43.5% each, i.e., a total by 87% [28]. We registered a majority rate of genotype E which corroborates the results found by Oluyinka and al. in 2015 Nigeria where all 72 cases of OBI were of genotype E, and Yousif and al., in 2014 in Sudan with 21.6% of genotype E. Furthermore in 2004, Mulders and al. in a multicenter study concerning the countries of West Africa, had already proved the existence of genotype A and E of HBV in Burkina Faso [29]. Our study confirms the presence of the E genotype in the context of occult HBV infection with a high prevalence of the E genotype. The E genotype of the HBV has a low genetic diversity according to Mulders and al., and there is no difference between the genotype E of HBV infection and that of occult HBV infection [24]. The A3 sub genotype registered in the present study is a genotype that is usually present in Central Africa (Gabon, Cameroon, D.R Congo and Central Africa) [30]; it has already been described in 2004 in Burkina Faso [29]. The persistence of this genotype could be explained by the migration flow of populations between the different areas of West and Central Africa. However, OBI genotypes A have been found in other parts of the world [9], thus Escobedo and al., at 2014 in Mexico registered a case among 24 children suffering from hepatitis, Youssif and al., at 2014 in Sudan registered a rate of 18.9%. We did not register cases of occult B infection with genotypes B, C, D, and F in the present study. While Hong Lin and al., in 2016 registered only genotypes B, C, and D in Chinese blood donors with OBI [31]. The absence of these genotypes in our context, could be explained not only by the geographic distribution of HBV infection, but also by the fact that we have not specifically targeted groups at high risk of OBI: chronic hepatitis patients, HCC patients, HCV/HIV co-infected, hemodialysis patients and immunocompromised patients. Indeed, a high prevalence of genotypes B (24% or 6/25) and D (32% or 8/25) was observed in HCC patients with OBI in Egypt [32] and these two genotypes were associated to a more rapid progression of the infection to HCC. In Saudi Arabia, genotype D was the most prevalent in occult B cases in blood donors (88.2% or 15/17). In our study we registered 5 cases (5/30 or 16.7%) of OBI recombination between genotypes A3 and E. Similar results have been reported by other authors. Thus, Makuwa and al, in 2006 in Gabon found 1 out of 13 cases of recombination between the A3 and E genotypes. Kurbanov and al., in 2005 also recorded cases of A3/E recombination (43.5%) in Cameroon. However, in Sudan, the recombinations in cases of Occult B infection mainly concerned the E/D genotypes with 13.5% [33].

We recorded 80% of cases of OBI seropositive against 20% of seronegativity to the antibody anti-HBc. These data corroborate those found by Makvandi and al. in Iran during a state of place on OBI (9). However, whether the OBI is seropositive or seronegative, the difference in the clinical plan always remains encrypted [34]. The anti-HBs protective marker was present in cases of OBI up to 10%, this phenomenon has already been described by Pernice and al. in Marburg in Germany. In fact, they noticed that the appearance of detectable HBsAc in the serum of patients, was correlated with the decrease or total absence of HBsAg [35]. Other authors have been able to highlight the appearance of the HBsAg/anti-HBs immune complex among patients of hepatocellular carcinoma with OBI [36].

## 5. Conclusion

At the end of this study, we were able to estimate a prevalence of 11.9% of cases of occult infection of the hepatitis B virus (OBI) in a population of 252 volunteers. After genotyping of cases of occult infections, genotypes B, C, D and F were absent while genotypes A3 and E were present with 23.3% and 60.0% respectively. A mixed infection with both E/A3 genotypes (16.7%) was also registered. OBI occupies a prominent place in certain groups of people such as blood donors, chronic hepatitis from HCV, PV/HIV and cancer patients. It would be linked to a much more rapid evolution of hepatitis towards hepatocellular carcinoma. The management of these groups of people must systematically include the search for viral DNA and/or anti-HBc antibodies in hepatocytes and/or in serum. However, the absence of circulating genotypes B and D of OBI cases constitutes a positive predictive value since their presence has been associated with a more rapid progression of the infection to hepatocellular carcinoma.

## Data Availability

All relevant data are within the manuscript and its Supporting Information files.

## 6. Acknowledgment

The research team thanks all the staff of Laboratory of Biology and Molecular Genetics (LAOBIGENE) of the University Ouaga I Pr Joseph Ki-Zerbo Burkina Faso and the Medical Biology Laboratory Department of the National Institute of Public Health/Bamako/Mali and all candidates who participated in the study.

## 7. Author contribution statement

Doumbia B and Simpore J, designed the study, analyzed and interpreted the results, and wrote the manuscript. Doumbia B, Diarra B, provided care for study participants, developed the registry, and reviewed the manuscript. Djigma F collected and checked the data, analyzed the data and reviewed the manuscript. Kone B and Baya B, developed an analytical approach for data analysis and interpretation and revised the manuscript. All authors have approved the final version of the manuscript before submission, so they are fully responsible for its content.

## References

1. World Health Organization. Hepatitis B Fact Sheet N204. Hepatitis B. World Health Organization. [Internet]. Available through: https://www.who.int/news-room/fact-sheets/detail/hepatitis-b

2. Tao I, Compaoré TR, Diarra B, Djigma F, Zohoncon TM, Assih M, and al. Seroepidemiology of hepatitis B and C viruses in the general population of burkina faso. Hepat Res Treat. 2014;2014:781843.

3. Simpore J, Savadogo A, Ilboudo D, Nadambega MC, Esposito M, Yara J, and al. Toxoplasma gondii, HCV, and HBV seroprevalence and co-infection among HIV-positive and -negative pregnant women in Burkina Faso. J Med Virol. June 2006;78(6):730–3.

4. Collenberg E, Ouedraogo T, Ganamé J, Fickenscher H, Kynast-Wolf G, Becher H, and al. Seroprevalence of six different viruses among pregnant women and blood donors in rural and urban Burkina Faso: A comparative analysis. J Med Virol. May 2006;78(5):683–92.

5. Tao I, Bisseye C, Nagalo BM, Sanou M, Kiba A, Surat G, and al. Screening of Hepatitis G and Epstein-Barr Viruses Among Voluntary non Remunerated Blood Donors (VNRBD) in Burkina Faso, West Africa. Mediterr J Hematol Infect Dis. 2013;5(1):e2013053.

6. Candotti D, Allain J-P. Transfusion-transmitted hepatitis B virus infection. J Hepatol. Oct. 2009;51(4):798–809.

7. Allain J-P, Candotti D. Diagnostic algorithm for HBV safe transfusion. Blood Transfus Trasfus Sangue. July 2009;7(3):174–82.

8. González R, Torres P, Castro E, Barbolla L, Candotti D, Koppelman M, and al. Efficacy of hepatitis B virus (HBV) DNA screening and characterization of acute and occult HBV infections among blood donors from Madrid, Spain. Transfusion (Paris). January 2010;50(1):221–30.

9. Makvandi M. Update on occult hepatitis B virus infection. World J Gastroenterol. 21 Oct. 2016;22(39):8720–34.

10. Somda KS, Sermé AK, Coulibaly A, Cissé K, Sawadogo A, Sombié AR, and al. Hepatitis B Surface Antigen Should Not Be the Only Sought Marker to Distinguish Blood Donors towards Hepatitis B Virus Infection in High Prevalence Area. Open J Gastroenterol. 9 Nov. 2016;6(11):362–72.

11. Samal J, Kandpal M, Vivekanandan P. Molecular mechanisms underlying occult hepatitis B virus infection. Clin MicroOBIl Rev. Jan. 2012;25(1):142–63.

12. Chaudhuri V, Tayal R, Nayak B, Acharya SK, Panda SK. Occult hepatitis B virus infection in chronic liver disease: full-length genome and analysis of mutant surface promoter. Gastroenterology. Nov. 2004;127(5):1356–71.

13. Lledó JL, Fernández C, Gutiérrez ML, Ocaña S. Management of occult hepatitis B virus infection: an update for the clinician. World J Gastroenterol. 28 March. 2011;17(12):1563–8.

14. Gupta S, Singh S. Occult hepatitis B virus infection in ART-naive HIV-infected patients seen at a tertiary care centre in north India. BMC Infect Dis. 7 March. 2010; 10:53.

15. Coppola N, Onorato L, Pisaturo M, Macera M, Sagnelli C, Martini S, and al. Role of occult hepatitis B virus infection in chronic hepatitis C. World J Gastroenterol. 14 Nov. 2015;21(42):11931–40.

16. Cacciola I, Pollicino T, Squadrito G, Cerenzia G, Orlando ME, Raimondo G. Occult hepatitis B virus infection in patients with chronic hepatitis C liver disease. N Engl J Med. 1 July. 1999;341(1):22–6.

17. Vivekanandan P, Daniel HD-J, Kannangai R, Martinez-Murillo F, Torbenson M. Hepatitis B virus replication induces methylation of both host and viral DNA. J Virol. May 2010;84(9):4321–9.

18. Xu R, Zhang X, Zhang W, Fang Y, Zheng S, Yu X-F. Association of human APOBEC3 cytidine deaminases with the generation of hepatitis virus B x antigen mutants and hepatocellular carcinoma. Hepatol Baltim Md. Dec. 2007;46(6):1810–20.

19. Tatematsu K, Tanaka Y, Kurbanov F, Sugauchi F, Mano S, Maeshiro T, and al. A genetic variant of hepatitis B virus divergent from known human and ape genotypes isolated from a Japanese patient and provisionally assigned to new genotype J. J Virol. Oct. 2009;83(20):10538–47.

20. Ma Y, Ding Y, Juan F, Dou XG. Genotyping the hepatitis B virus with a fragment of the HBV DNA polymerase gene in Shenyang, China. Virol J. 22 June 2011;8:315.

21. Zhou J-Y, Zhang L, Li L, Gu G-Y, Zhou Y-H, Chen J-H. High hepatitis B virus load is associated with hepatocellular carcinomas development in Chinese chronic hepatitis B patients: a case control study. Virol J. 13 June 2012;9:16.

22. Ozaslan M, Ozaslan E, Barsgan A, Koruk M. Mutations in the S gene region of hepatitis B virus genotype D in Turkish patients. J Genet. Dec. 2007;86(3):195–201.

23. Yeh C-T, Chen T, Hsu C-W, Chen Y-C, Lai M-W, Liang K-H, and al. Emergence of the rtA181T/sW172* mutant increased the risk of hepatoma occurrence in patients with lamivudine-resistant chronic hepatitis B. BMC Cancer. 21 Sept. 2011;11:398.

24. Oluyinka OO, Tong HV, Bui Tien S, Fagbami AH, Adekanle O, Ojurongbe O, and al. Occult Hepatitis B Virus Infection in Nigerian Blood Donors and Hepatitis B Virus Transmission Risks. PloS One. 2015;10(7):e0131912.

25. Baghbanian M, Halvani M, Roghani HS, Lotfi MH, Yazdi MF. Prevalence of occult hepatitis b infection in iranian cancer patients before chemotherapy treatment. Arq Gastroenterol. Sept. 2016;53(3):175–9.

26. Banerjee A, Kurbanov F, Datta S, Chandra PK, Tanaka Y, Mizokami M, and al. Phylogenetic relatedness and genetic diversity of hepatitis B virus isolates in Eastern India. J Med Virol. Sept. 2006;78(9):1164–74.

27. Minuk GY, Sun D-F, Uhanova J, Zhang M, Caouette S, Nicolle LE, and al. Occult hepatitis B virus infection in a North American community-based population. J Hepatol. April 2005;42(4):480–5.

28. Kurbanov F, Tanaka Y, Fujiwara K, Sugauchi F, Mbanya D, Zekeng L, and al. A new subtype (subgenotype) Ac (A3) of hepatitis B virus and recombination between genotypes A and E in Cameroon. J Gen Virol. July 2005;86(Pt 7):2047–56.

29. Mulders MN, Venard V, Njayou M, Edorh AP, Bola Oyefolu AO, Kehinde MO, and al. Low genetic diversity despite hyperendemicity of hepatitis B virus genotype E throughout West Africa. J Infect Dis. 15 July 2004;190(2):400–8.

30. Makuwa M, Souquière S, Telfer P, Apetrei C, Vray M, Bedjabaga I, and al. Identification of hepatitis B virus subgenotype A3 in rural Gabon. J Med Virol. Sept. 2006;78(9):1175–84.

31. Lin H, Zhao H, Tang X, Hu W, Jiang N, Zhu S, and al. Serological Patterns and Molecular Characterization of Occult Hepatitis B Virus Infection among Blood Donors. Hepat Mon. Oct. 2016;16(10):e40492.

32. Hassan ZK, Hafez MM, Mansor TM, Zekri ARN. Occult HBV infection among Egyptian hepatocellular carcinoma patients. Virol J. 3 March 2011;8:90.

33. Yousif M, Mudawi H, Hussein W, Mukhtar M, Nemeri O, Glebe D, and al. Genotyping and virological characteristics of hepatitis B virus in HIV-infected individuals in Sudan. Int J Infect Dis IJID Off Publ Int Soc Infect Dis. Dec. 2014;29:125–32.

34. Torbenson M, Thomas DL. Occult hepatitis B. Lancet Infect Dis. August 2002;2(8):479–86.

35. Pernice W, Sodomann CP, Lüben G, Seiler FR, Sedlacek HH. Antigen-specific detection of HBsAG-containing immune complexes in the course of hepatitis B virus infection. Clin Exp Immunol. August 1979;37(2):376–80.

36. Brown SE, Howard CR, Steward MW, Ajdukiewicz AB, Whittle HC. Hepatitis B surface antigen containing immune complexes occur in seronegative hepatocellular carcinoma patients. Clin Exp Immunol. Feb. 1984;55(2):355–9.

